# Accuracy and reliability of an MR-compatible dorsiflexion ergometer for dynamic 31P-MRS: Comparison with a clinical dynamometer in individuals with and without obesity

**DOI:** 10.1101/2025.04.13.25325753

**Authors:** Anders Stouge, Ole Emil Andersen, Esben Søvsø Szocska Hansen, Cuno Rasmussen, Ryan Godsk Larsen, Gwenael Layec, Jack Miller, Ladislav Valkovič, Jane A. Kent, Christoffer Laustsen, Jens Meldgaard Bruun, Henning Andersen, Michael Vaeggemose

**Affiliations:** The MR Research Centre, Aarhus University, Aarhus, Denmark; Steno Diabetes Center Aarhus, Aarhus, Denmark; Department of Neurology, Aarhus University Hospital, Aarhus, Denmark; Department of Clinical Medicine, Aarhus University, Aarhus, Denmark; Department of Public Health, Section of Sport science, Aarhus University, Aarhus, Denmark; Department of Clinical Pharmacology, Aarhus University Hospital, Aarhus, Denmark; ExerciseTech, Department of Health Science and Technology, Aalborg University, Aalborg, Denmark; School of Health and Kinesiology, University of Nebraska Omaha, NE, USA; Clarendon Laboratory, Department of Physics, University of Oxford, Oxford, United Kingdom; Oxford Centre for Clinical Magnetic Resonance Research (OCMR), Division of Cardiovascular Medicine, Radcliffe Department of Medicine, University of Oxford, Oxford, United Kingdom; Department of Imaging Methods, Institute of Measurement Science, Slovak Academy of Sciences, Bratislava, Slovakia; Muscle Physiology Laboratory, Department of Kinesiology, University of Massachusetts, Amherst, MA, USA; GE HealthCare, Brondby, Denmark

**Author notes:** Corresponding author: Anders Stouge, MD (AS).

## Abstract

**Background:** The clinical application of MR-compatible ergometers for muscle contractile assessment is limited by a lack of validation against standard clinical dynamometers. Moreover, the impact of obesity on the reliability of MR ergometer-based muscle contractile assessments and the quality of phosphorus-31 magnetic resonance spectroscopy (^31^P-MRS) data remains unclear. This study aimed to validate an MR-compatible ergometer against a clinical dynamometer and to evaluate the applicability of ^31^P-MRS in individuals with severe obesity.

**Methods:** Twenty adults (35–60 years) were recruited and divided into groups of non-obese (BMI 18.5-30 kg/m^2^, n=10) and severely obese (BMI ≥35 kg/m², n=10), matched for age, sex, and height. Ankle dorsiflexion was assessed using both a clinical dynamometer and an MR ergometer, measuring maximal voluntary isometric contraction (MVIC) and a 4-minute isotonic fatiguing exercise. ^31^P-MRS was continuously acquired during the in-scanner exercise. Agreement between devices was assessed using Bland-Altman plots and intraclass correlation coefficients (ICCs). ^31^P-MRS data quality was evaluated based on signal-to-noise ratio (SNR), uncertainty of fit (CRLB), and phosphocreatine (PCr) recovery fit (R^2^). Pearson’s correlations examined relationships between muscle fatigue and metabolic parameters.

**Results:** All subjects successfully completed the protocol on both devices. The MR ergometer demonstrated moderate-to-excellent reliability (ICC ≥0.50) for most contractile parameters. While maximal torque, power, and work were underestimated on the MR ergometer (16-28%), this bias was consistent across BMI groups. ^31^P-MRS met preset quality thresholds (SNR≥5, CRLB <20%, R^2^≥0.70) in both groups. Dorsiflexion fatigue (reduction in power) correlated strongly (r ≥0.77) with metabolic changes, including PCr depletion (R²=0.68), pH drop (R²=0.59), PCr recovery time constant (R²=0.62), and inorganic phosphate accumulation (Pi/PCr) (R²=0.67).

**Conclusion:** The MR ergometer demonstrated feasibility, acceptable reliability, and consistent ^31^P-MRS data quality across BMI groups. These findings support the use of the MR ergometer for in-scanner dorsiflexor assessments, even in individuals with severe obesity.

## Introduction

Phosphorus (^31^P) magnetic resonance spectroscopy (MRS) is a unique non-invasive modality for assessing skeletal muscle metabolism [1]. In contrast to invasive muscle biopsies, ^31^P-MRS is able to monitor in-vivo metabolic changes with high temporal resolution in skeletal muscle during contractile activity, and subsequent recovery [2]. Moreover, ^31^P-MRS is a well-established tool for evaluating muscle and mitochondrial health; key factors in the pathophysiology of conditions such as obesity and type 2 diabetes [3]. Given the rising prevalence of these conditions as a major public health concern [4, 5], future large scale clinical trials incorporating ^31^P-MRS are warranted to develop strategies to mitigate functional decline and promote healthy aging.

However, applying ^31^P-MRS in clinical populations, particularly in individuals with obesity, presents several technical challenges. A key challenge is the reliance on MR-compatible ergometers for in-scanner exercise, which are often custom-built, lack standardization, and validation against clinically accepted dynamometers [6]. Furthermore, obesity introduces practical constraints, including difficulties in achieving proper positioning for in-scanner muscle assessment due to anthropometric factors. Excess adipose tissue can further compromise data quality by increasing signal attenuation, field inhomogeneity, and introducing motion artifacts, making muscle metabolic assessments more challenging [7]. Consequently, individuals with severe obesity (BMI ≥ 35 kg/m^2^) remain underrepresented in ^31^P-MRS research, even in studies examining populations with excess adipose tissue [8–13]. This underrepresentation limits the exploration of this high-risk group and reduces the generalizability of findings.

To address these challenges, efforts have been made to develop MR-compatible exercise devices that are cost-effective, biomechanically precise, and support multiple exercise modalities while emphasizing clinical applicability [14–17]. However, further advancements are needed to validate these devices for broader clinical adoption, particularly in individuals with obesity. A crucial next step is to establish performance equivalence between MR-compatible ergometers and clinically validated dynamometers. An initiative that, to our knowledge, has not yet been undertaken.

Given these challenges, this study aimed to validate a custom-built MR-compatible ergometer by comparing its performance to a clinically validated dynamometer for muscle contractile assessment [18–20]. Specifically, we aimed to: (1) evaluate the feasibility and reliability of in-scanner dorsiflexion assessments in individuals with normal-to-overweight BMI (BMI 18.5-30 kg/m^2^) and those with class II or III obesity (BMI ≥ 35 kg/m^2^), and (2) provide proof-of-concept for ^31^P-MRS in severe obesity by assessing spectral quality and examining whether the interrelation between ^31^P-MRS parameters and muscle contractile properties remained strong and consistent, supporting its applicability for muscle function evaluation in this population.

## Materials and Methods

### MR ergometer description

The MR-compatible ergometer was constructed using non-magnetic materials and engineered to operate within a standard 3 Tesla whole-body MR imaging system. A schematic representation of the MR ergometer setup is provided in Figure 1. Detailed construction specifications are provided in supporting information S1. All essential components required for constructing the MR ergometer are accessible for 3D printing via Onshape (Onshape, PTC Inc.; link is available upon reasonable request).

**Fig 1.**
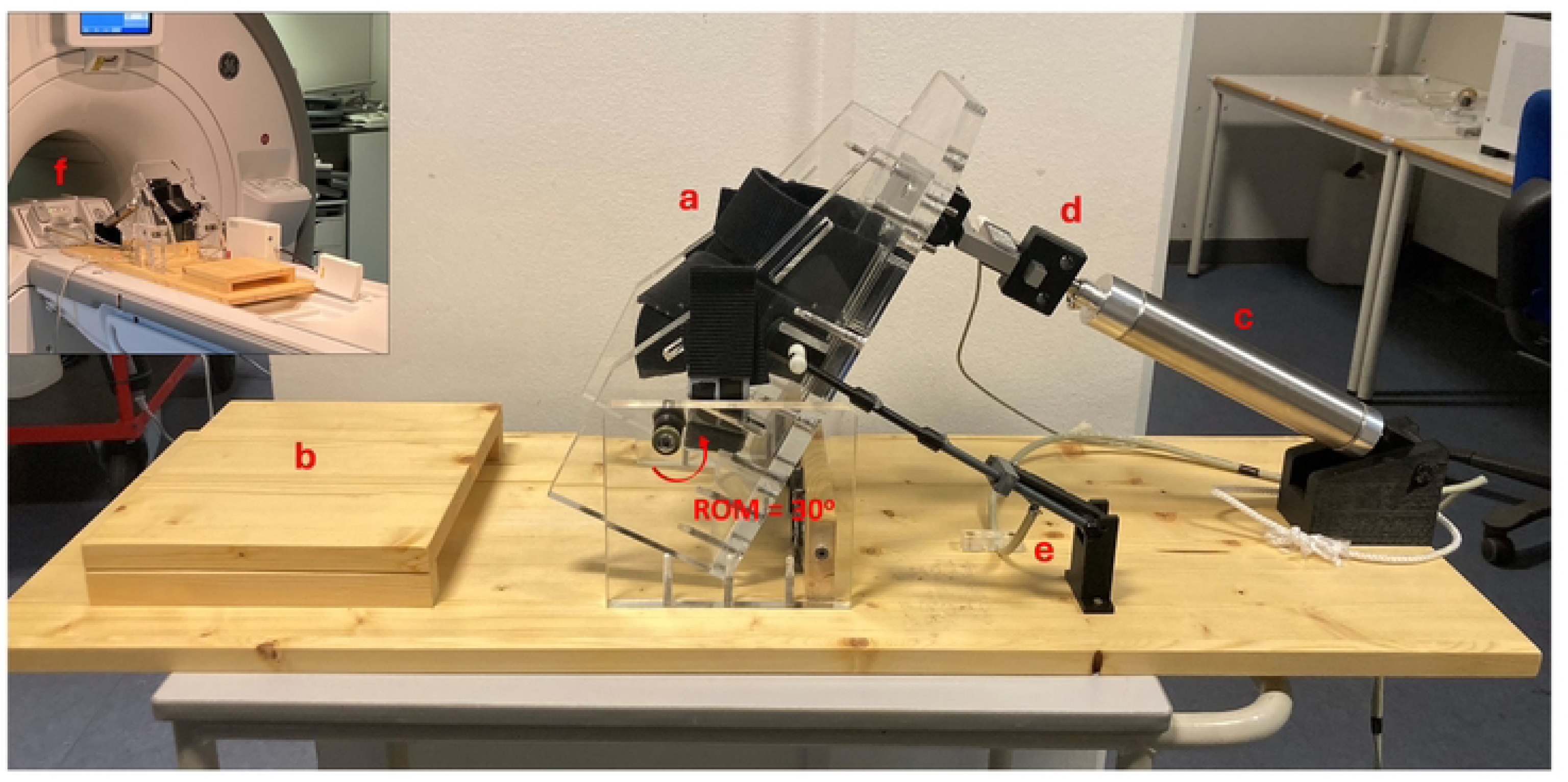
The MR ergometer. (a) Pedal with hook-and-loop fasteners for foot fixation, (b) Calf support, (c) Pneumatic cylinder for resistance control, (d) Strain gauge, (e) Potentiometer, (f) Adapter plate fixation to the MR table. ROM = Range of Motion.

The MR ergometer features a footplate supporting isotonic dorsiflexion, plantarflexion, and isometric exercise, allowing ankle joint rotation from 30° to 0° plantarflexion. Foot placement was secured using two hook-and-loop fastener straps across dorsum of the foot. The ankle’s axis of rotation was adjusted via a hook-and-loop heel support, which also prevented the foot from slipping down the plate (Fig. 1a). A custom-built plantar flexor support plate was affixed to the adapter plate to further ensure proper alignment of the axis of rotation and enhance subject comfort during exercise (Fig. 1b). Resistance was controlled by a pneumatic cylinder (Model 176-DX, Bimba, IL, USA) (Fig. 1c), with workload adjustments modulated by a dedicated pneumatic regulator system (S1 Fig. 2). Pedal movement and applied force were recorded based on a potentiometer (Slide-010KA-Stereo, Philips, Amsterdam, Netherlands) and strain-gauge (Model SSM/SSM2, Interface, Scottsdale, AZ, USA), respectively (Fig. 1d+e). Position and torque data were sampled at 1000 Hz (NI-DAQ USB-6008, National Instruments, TX, USA) and displayed in real time in the MR control room using an in-house analysis program (Footlogger1.1, Department of Public Health, Section of Sport Science, Aarhus University, Denmark) (S1 Fig. 1a). Acquired data from exercise protocols was processed at 100 Hz using a custom MATLAB script (MATLAB R2022b, MathWorks, Natick, MA, USA) (Fig. 2c).

**Fig 2.**
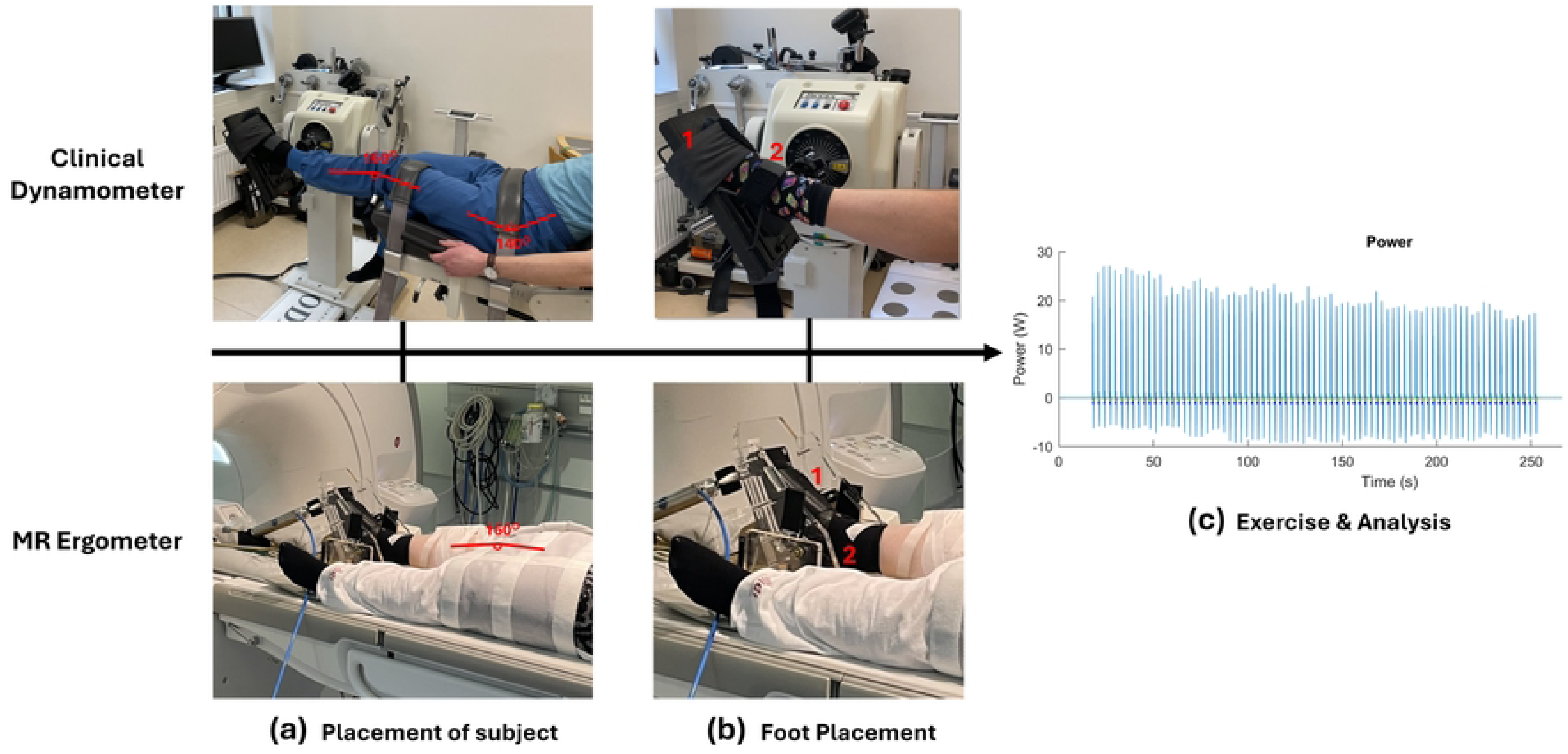
Exercise protocol setup on the clinical dynamometer and MR ergometer. (a) Subject positioning on the Biodex System 3 Pro (top) and MR ergometer (bottom). (b) Foot fixation to the footplate in both setups: (1) Hook-and-loop fasteners for foot fixation. (2) Rotational axis of the foot. (c) Exercise protocol and data processing using a unified analysis pipeline for both setups.

### Subjects and exercise protocol

Two groups of ten subjects were enrolled in the study between December 7, 2022, and November 22, 2024 (Table 1). The Non-obese (Non-O) group consisted of individuals with normal weight or overweight (BMI 18.5-30 kg/m^2^), while the Obesity (O) group included individuals with class II or III obesity (BMI ≥ 35 kg/m^2^). The groups were matched on age, sex and height, and all subjects were sedentary to recreational active. Written informed consent was obtained from all subjects in accordance with the requirements of the Danish research ethics committee.

**Table 1.**
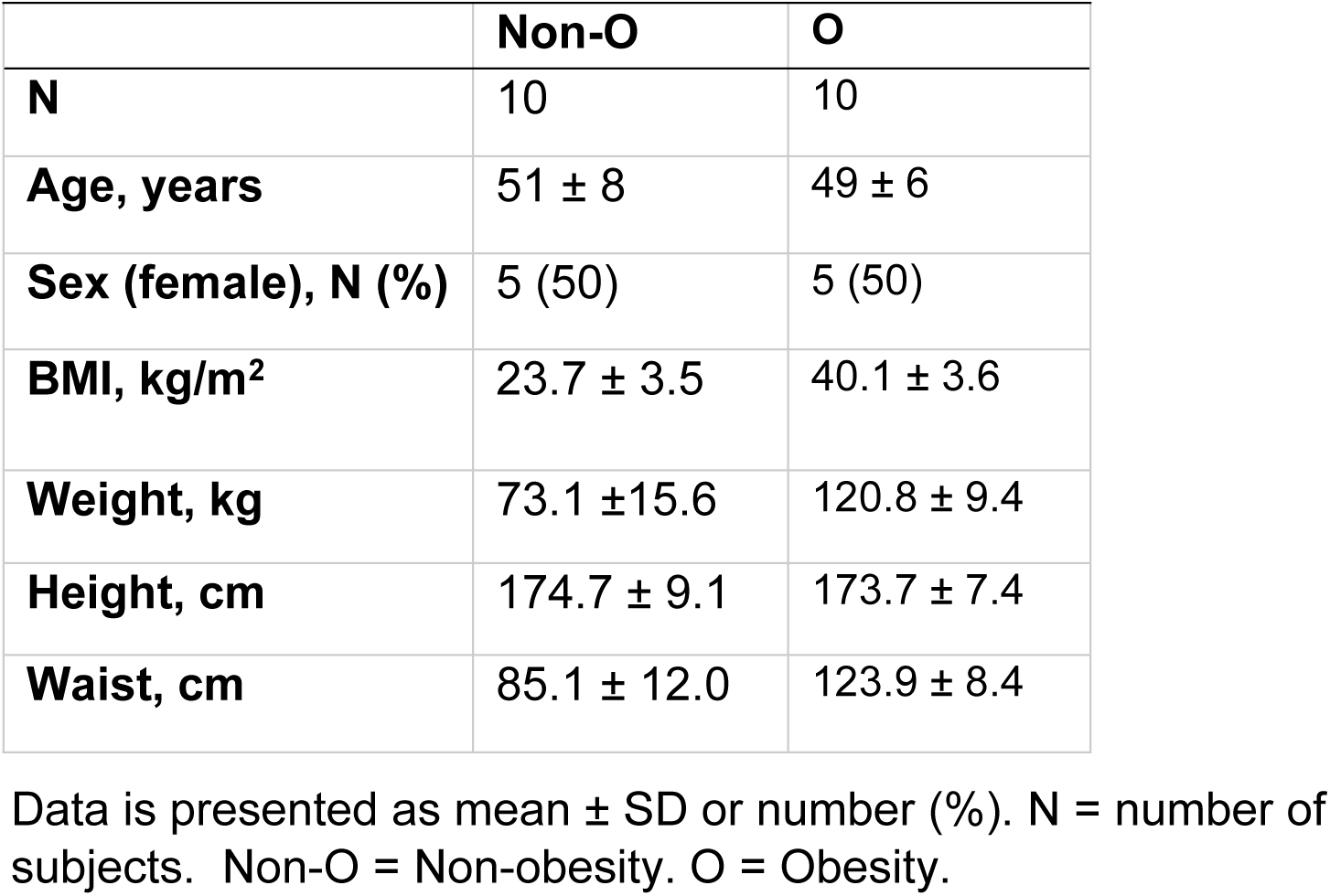
Clinical characteristics of study groups.

All subjects completed the exercise protocol on both a Biodex System 3 Pro dynamometer ® (Biodex Medical Systems, NY, USA) and on the MR-compatible ergometer (Fig. 2). The clinical dynamometer exercise session was conducted first, followed by the MR session, with a duration of three to seven days between sessions. Prior to the main exercise protocol, subjects completed a warm-up session on both devices to familiarize themselves with the setup. Moreover, to determine maximal dorsiflexion strength, subjects performed three Maximal Voluntary Isometric Contractions (MVICs), spaced 45 seconds apart, ten minutes before initiating the isotonic exercise protocol. MVIC was defined as the highest peak torque (Nm) achieved in the series, following the standard procedure previously outlined in our lab [18]. The isotonic exercise protocol involved four minutes of concentric dorsiflexion, with contractions performed every three seconds cued by a metronome. Resistance was set to 30% of the subject’s MVIC, and subjects were instructed to perform every contraction ‘as hard and fast as possible’. Verbal encouragement was given and standardized across both setups. In all cases the exercise was performed with the nondominant leg.

#### Clinical dynamometer setup

Subjects were placed in a supine position with hip and knee angles of 140° and 160°, respectively (Fig. 2a). These joint angles were selected to approximate the position used on the second visit for the exercise protocol on the MR ergometer. To isolate ankle dorsiflexion, a strap was placed across the thigh and hip. The foot was firmly fixed to the footplate using a hoop-and-loop fastener positioned across the dorsum of the foot (Fig. 2b). The rotational axis of the footplate was aligned just below and posterior to the medial malleolus (Fig. 2b). All contractions were performed within a 30° range of motion (ROM), from 30° to 0° plantarflexion. This ROM was consistent with the MR ergometer setup and fell within the physiological range of all subjects.

#### MR ergometer setup

Subjects were positioned supine, with both hip and knee joints set at approximately 160° angles (Fig. 2a). A strap was placed across the thigh to isolate ankle dorsiflexion, and the foot was secured similar to the clinical dynamometer setup (Fig. 2b), as described previously for the MR ergometer description.

Muscle power and work were computed, during the concentric phase of each contraction, as the product of torque and velocity, and torque and the acquired range of motion, respectively. Initial values for maximum torque, maximum power, maximum angular velocity, and work were quantified as an average derived from the first five contractions. Muscle fatigue was assessed as the relative decline in power and work, expressed as a percentage drop from the first five contractions to the last five contractions.

### ^31^P-MRS acquisition and processing

^31^P-MRS data was acquired on a whole-body 3T MR scanner (MR750, GE Healthcare, Waukesha, WI, USA) with a bore size of 60cm, equipped with a dual-channel ^1^H/^31^P surface coil tuned to 51.72 MHz and 127.7 MHz (RAPID Biomedical GmbH, Rimpar, Germany). The dimensions of the coil were 4 x 3 cm for ^31^P and 8 x 8 cm for ^1^H, with a housing size of 9 x 15 cm. The coil was fixed on the shin using medical silicone tape, with proximal border of the coil housing aligned horizontally with the tibial tuberosity and the head of the fibula. The lateral border of the coil housing was positioned along the length of the anterior tibial crest.

Scout images were used to verify placement of the surface coil over the tibialis anterior muscle prior to the phosphorus acquisition (Fig. 3a). Transmission power and center frequency were calibrated to optimize the phosphorus acquisition using a Bloch-Siegert off-resonance approach [21]. ^31^P-MRS were acquired unlocalized using a hard pulse (500 μs) with the following parameters: Repetition time (TR) = 2s, effective flip angle (FA) = 70°, bandwidth = 5 kHz, and 1024 complex points. The length of the protocol was 16 minutes, consisting of 2 minutes of rest, 4 minutes of exercise, and 10 minutes of recovery. The workflow for ^31^P-MRS data processing is illustrated in Figure 3. Spectra averaging was adapted from Fitzgerald et al. (2023) [22], achieving temporal resolutions of 90 s during rest (with the first 30 s discarded as dummy pulses) and 10 s during exercise. For the recovery period, spectral resolutions were set to 4 s for the first 20 s, 8 s for the next 280 s, and 30 s for the remaining 5 min (see Fig. 3f for an example of the temporal data). All free induction decays (FIDs) were processed using a custom-built MATLAB script that incorporated: (1) the open-source MRS analysis toolbox OXSA, based on the work of Purvis et al. (2017), for Advanced Method for Accurate, Robust and Efficient Spectral (AMARES) fitting of MRS data [23], and (2) a spectral analysis pipeline adapted from Bartlett et al. [24]. Pre-processing steps included DC offset correction and zero-filling to 2048 points before Fourier transformation (Fig. 3c). AMARES fitting quantified area under the peak of phosphomonoesters (PME), inorganic phosphate (Pi), phosphodiesters (PDE), phosphocreatine (PCr), and the α-, β- and γ-phosphate groups of ATP (Fig. 3d+g). Intracellular pH was calculated based on the chemical shift of Pi relative to PCr [25]:

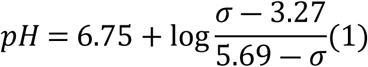

The parameter σ represents the chemical shift of Pi relative to PCr, expressed in parts per million (ppm). Broadening and splitting of the Pi peak may occur during exercise, reflecting the emergence of two distinct pH pools within the muscle [26]. In cases where Pi splitting was observed, the pH corresponding to each Pi pool was calculated individually (Fig. 3i), and a weighted average, based on the total amplitude of the Pi peaks, was then used to estimate the overall pH [27]:

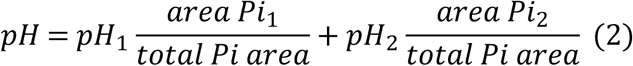

PCr resynthesis during the 10-minute recovery period was modeled using a bi-exponential function to determine the rate constant (k_PCr_) [28] (Fig. 3i):

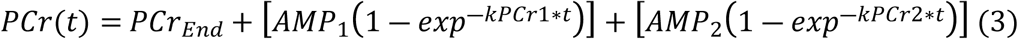

Where PCr(t) represents the calculated [PCr] at time point t, and PCr_end_ is the measured [PCr] at the end of the four minutes of exhaustive exercise. AMP_1_ and AMP_2_ correspond to the amplitudes of the primary and secondary recovery components (in mM), and k_PCr1_ and k_PCr2_ are their respective rate constants. To prevent overfitting and ensure physiologically realistic parameter estimations, the sum of AMP_1_ and AMP_2_ was constrained to not exceed the difference between [PCr]_end_ and [PCr]_rest_. The PCr recovery time constant (τ_PCr_) was derived as the reciprocal of k_PCr_.

**Fig 3.**
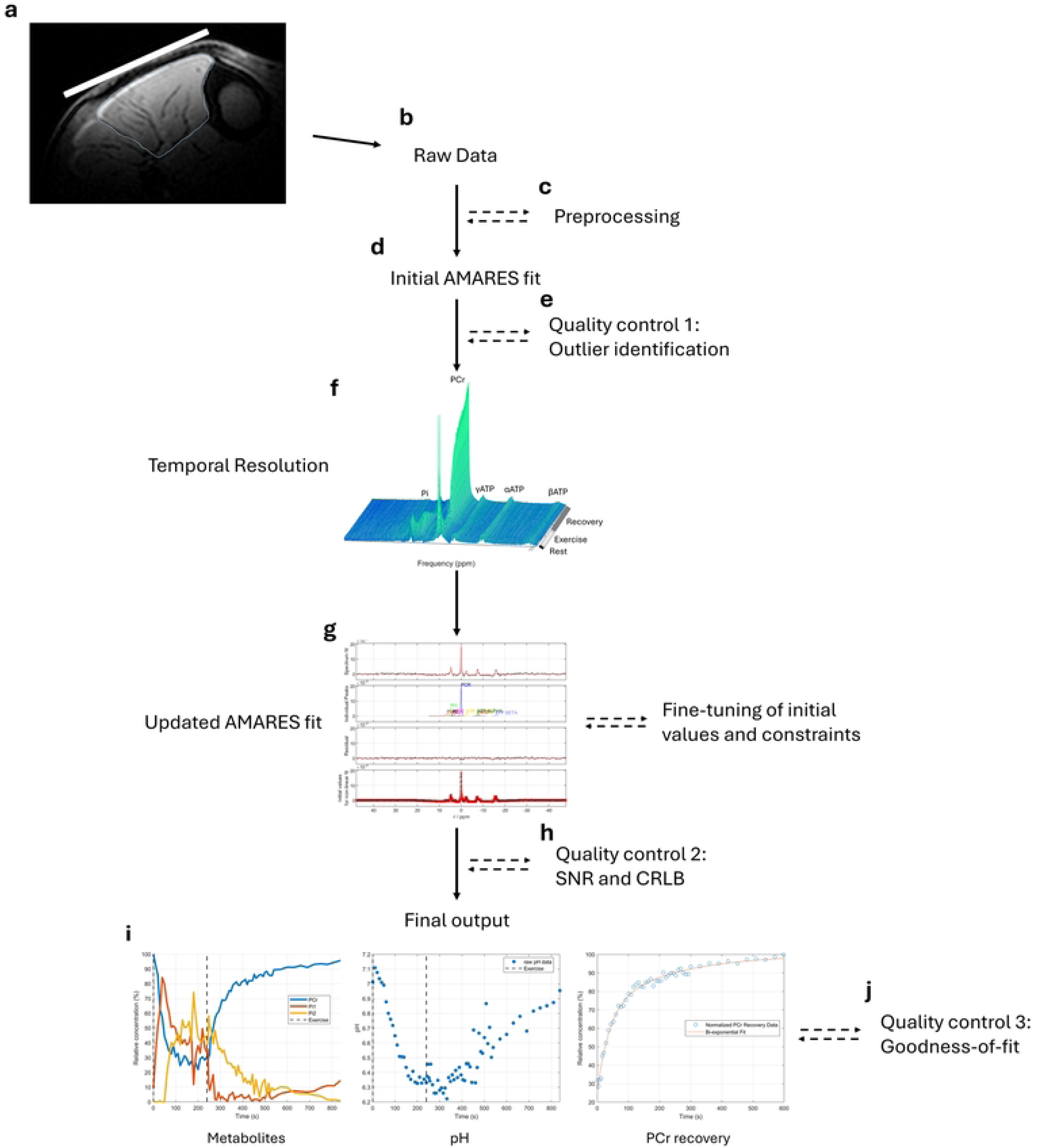
Flowchart of 31P-MRS data processing. a) <u>Coil placement</u>: Illustration of coil placement (white rectangle) and the region of interest dorsiflexors, dotted line) for an Obesity (O) individual. b) <u>Raw data acquisition</u>: Data analysis begins with loading raw FID files. c) <u>Preprocessing:</u> DC offset correction and zero-filling applied before the initial AMARES itting. d) <u>AMARES fitting</u>: Performed on the preprocessed spectra. e) <u>Quality control 1:</u> Outlier detection based on the AMARES fit of preprocessed data. f) <u>Temporal resolution:</u> Temporal visualization of all averaged spectra. g) <u>Refined AMARES fitting:</u> Averaged spectra are reanalyzed using an updated prior-nowledge file based on individual dataset. This step can be iterated if further improvement is eeded. h) <u>Quality control 2:</u> Evaluation of Signal-to-Noise Ratio (SNR) and Cramér-Rao Lower Bound (CRLB) focusing on rest and end-exercise conditions. i) <u>Final output:</u> Extracted dynamic metabolite concentrations, pH, and PCr recovery fit. j) <u>Quality control 3:</u> Goodness-of-fit assessment for PCr recovery kinetics. Solid arrows indicate the main workflow, while dashed arrows represent optional processing teps.

The data quality check pipeline for ^31^P-MRS data was adapted from recent work by Naëgel et al. [29] and Xu et al. [30], and consisted of the following steps:

1. <u>Outlier identification</u>: Outliers in the raw pre-processed data were detected based on the time derivative sum of PCr, Pi, and γATP (Fig. 3e). An outlier was defined as a data point deviating by more than four standard deviations (SD) from a three-point moving mean and was removed from further analysis. Theoretically, large variations in the sum of PCr + Pi + γATP may indicate corrupted data (e.g., movement artifacts) since this sum of phosphates should remain relatively constant during exercise [31].
2. <u>Signal-to-Noise Ratio (SNR):</u> SNR was assessed for PCr, Pi, and γATP at rest and at the end of exercise. SNR was calculated as the fitted amplitude divided by the SD of the noise, derived from the AMARES fitting results [32], with an SNR ≥ 5 considered acceptable (Fig. 3h).
3. <u>Cramér-Rao Lower Bound (CRLB</u>): The uncertainty of fit was evaluated using CRLB, expressed as a percentage, for PCr, Pi, and γATP at rest and end-exercise. CRLB was computed from AMARES fitting results, incorporating the linear relationship between parameters provided by prior knowledge to ensure robust evaluation [30]. A CRLB < 20% was considered acceptable (Fig. 3h).
4. <u>Goodness-of-fit for PCr recovery</u>: The fit quality of PCr recovery was evaluated using the coefficient of determination (R^2^) (Fig. 3j). A coefficient of determination ≥ 70% was considered acceptable [29].

### Concepts and definitions for method comparison

In this study, reproducibility and reliability are used interchangeably due to the method comparison design, examining variation in muscle contractile parameters between the two modalities and the accuracy of the MR ergometer outputs [33]. Validity refers to how well measured muscle contractile parameters and ^31^P-MRS reflect the intended physiological constructs. This study considers construct validity (e.g. whether our spectral fitting accurately quantified ^31^P-MRS signals) and criterion validity (e.g. the interrelation between muscle fatigue and ^31^P-MRS parameters) [34]. Feasibility refers to the overall ability to perform muscle contractile assessments on individuals with and without obesity using the MR ergometer.

### Statistical analysis

All statistical analyses were conducted using STATA 16 (StataCorp LCC, TX, USA). Data normality was assessed through visual inspection of Q-Q plots and histograms. The agreement between MR ergometer and clinical dynamometer was evaluated using Bland-Altman plots, to assess: 1) Heteroscedasticity. Whether the differences between measurements varied with the magnitude of the mean, 2) Systematic Bias. The presence of a mean difference between the two setups deviating from zero, 3) Random error. The variability of the differences between setups, calculated as the SD of the differences multiplied by 1.96 divided by the grand mean and expressed as a percentage. 4) Limits of Agreement (LOA). Indicating the absolute agreement between the two setups.

Heteroscedasticity was considered present if the R^2^ value exceeded 0.1 [35] and if this finding was further confirmed by visual inspection. For datasets exhibiting heteroscedasticity on absolute measurements, Bland-Altman plots using log-transformed data were created to perform relative scale comparisons. Reliability of the MR ergometer outputs was evaluated using the intraclass correlation coefficient (ICC), calculated with a two-way mixed-effects model, provided the data demonstrated homoscedasticity. A reliability threshold of ICC ≥

0.50 was set as the lower limit to suggest clinical usefulness [36]. Differences in systematic bias between groups for muscle MVIC, power and work parameters were analyzed using two-tailed Student’s t tests unless otherwise specified. The relationship between muscle fatigue and ^31^P-MRS measures was assessed using Pearson’s correlation. Group differences in the relationship of variables were tested using moderated regressions with an interaction term. All results are reported as mean ± SD, with statistical significance set at p-value < 0.05.

## Results

### Clinical dynamometer vs. MR ergometer

All subjects completed the exercise protocol on both the clinical dynamometer and MR ergometer. The agreement between measurements obtained from the clinical dynamometer and MR ergometer is summarized in Table 2. Consistency between the two methods, as assessed by ICC, indicated moderate-to-excellent reliability (≥0.50) for most parameters.

**Table 2.**
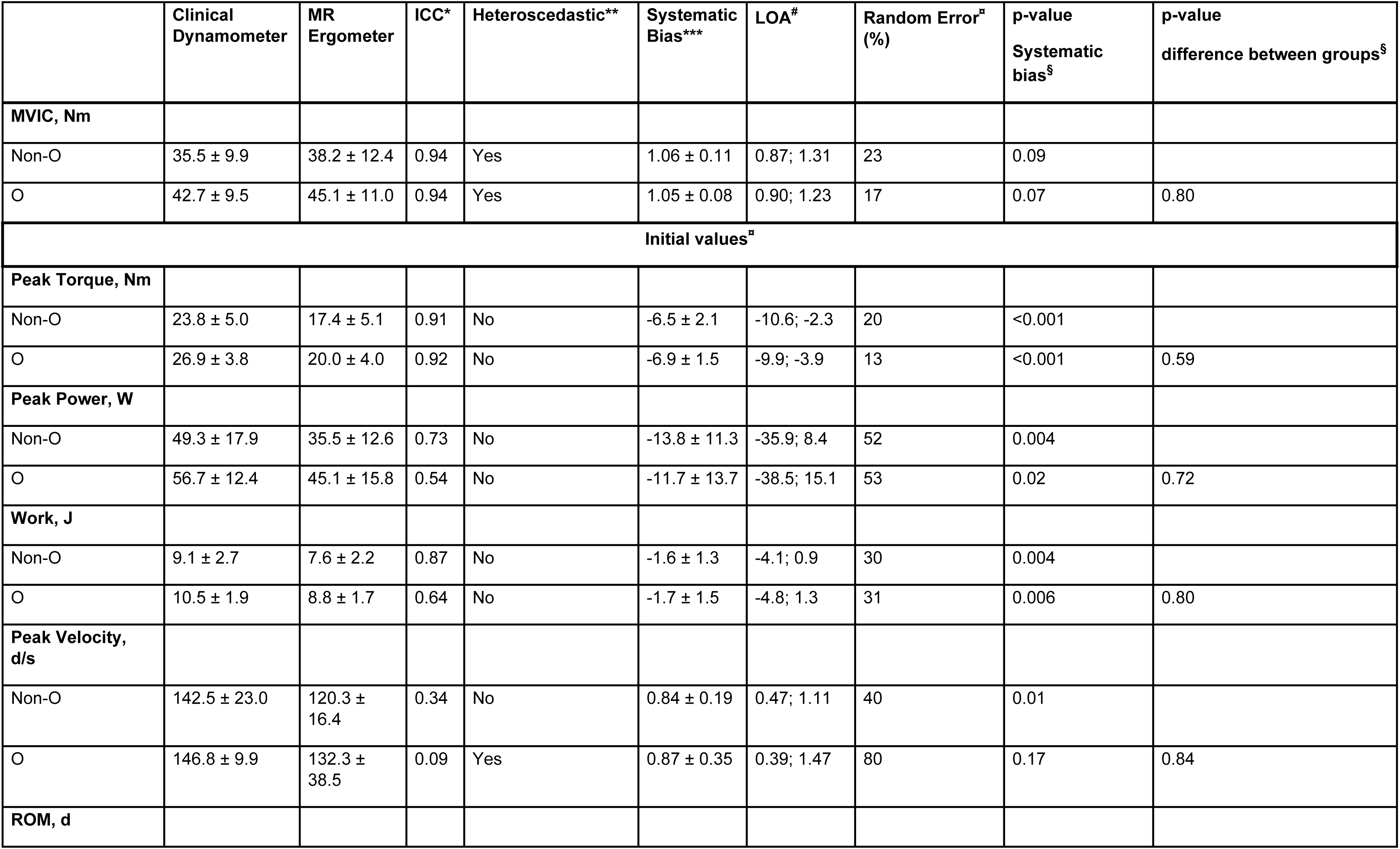
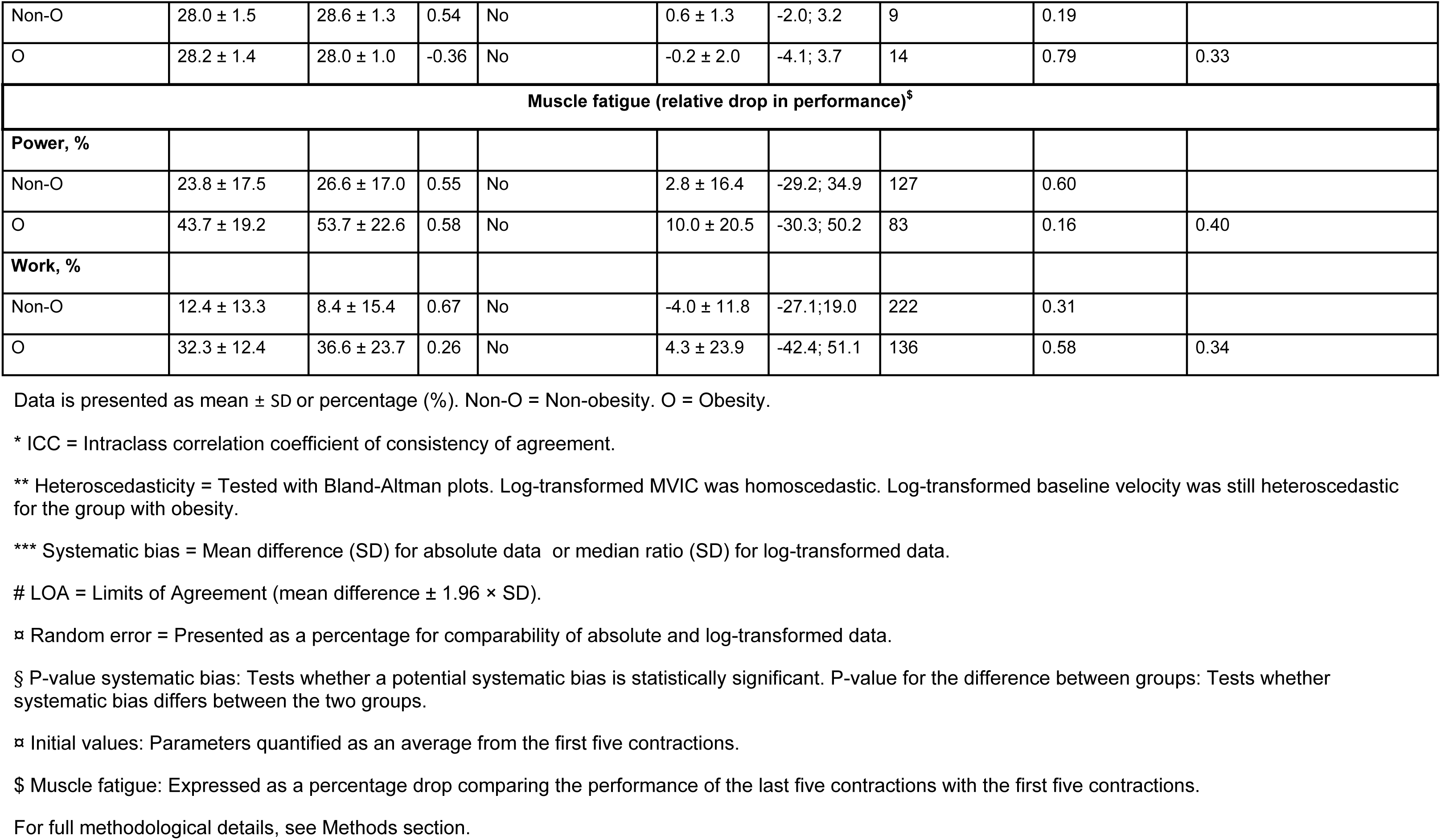
Reproducibility of ankle dorsiflexion measurements: Clinical dynamometer vs. MR ergometer.

However, initial peak velocity, initial ROM, and muscle fatigue (assessed as relative drop in muscle work) demonstrated lower reliability. Systematic bias was observed in the MR ergometer measurements for initial contractile values. Specifically, the MR ergometer underestimated initial peak torque (27% vs. 26%), peak power (28% vs. 21%), and work (18% vs. 16%) for Non-O and O groups, respectively, as well as initial peak velocity for the Non-O group (16%).However, the systematic differences between devices remained consistent across groups. No systematic bias was observed for MVIC or muscle fatigue parameters, though MVIC tended to be slightly overestimated on the MR ergometer (6 % for Non-O and 5% for O, P > 0.05). Random errors varied significantly, ranging from 9% to 222% for the Non-O group and 13% to 136% for the O group. Initial muscle work and its derivatives (peak torque and ROM) exhibited the lowest random errors (9% to 31%), compared to higher errors in velocity-dependent measures (peak power: 52 to 53%, and peak velocity: 40% to 80%). In contrast, for fatigue parameters, muscle work exhibited the highest random errors (136% to 222%), while power errors ranged from 83% to 127%. Heteroscedasticity was observed for absolute values of MVIC and initial peak velocity, while MVIC demonstrated homoscedasticity when analyzed on a relative scale. Bland-Altman plots for muscle contractile parameters are provided in supporting information S2.

### Data quality of ^31^P-MRS parameters

The dynamic changes in metabolite levels and pH during exercise are presented in Figure 4.

**Fig 4.**
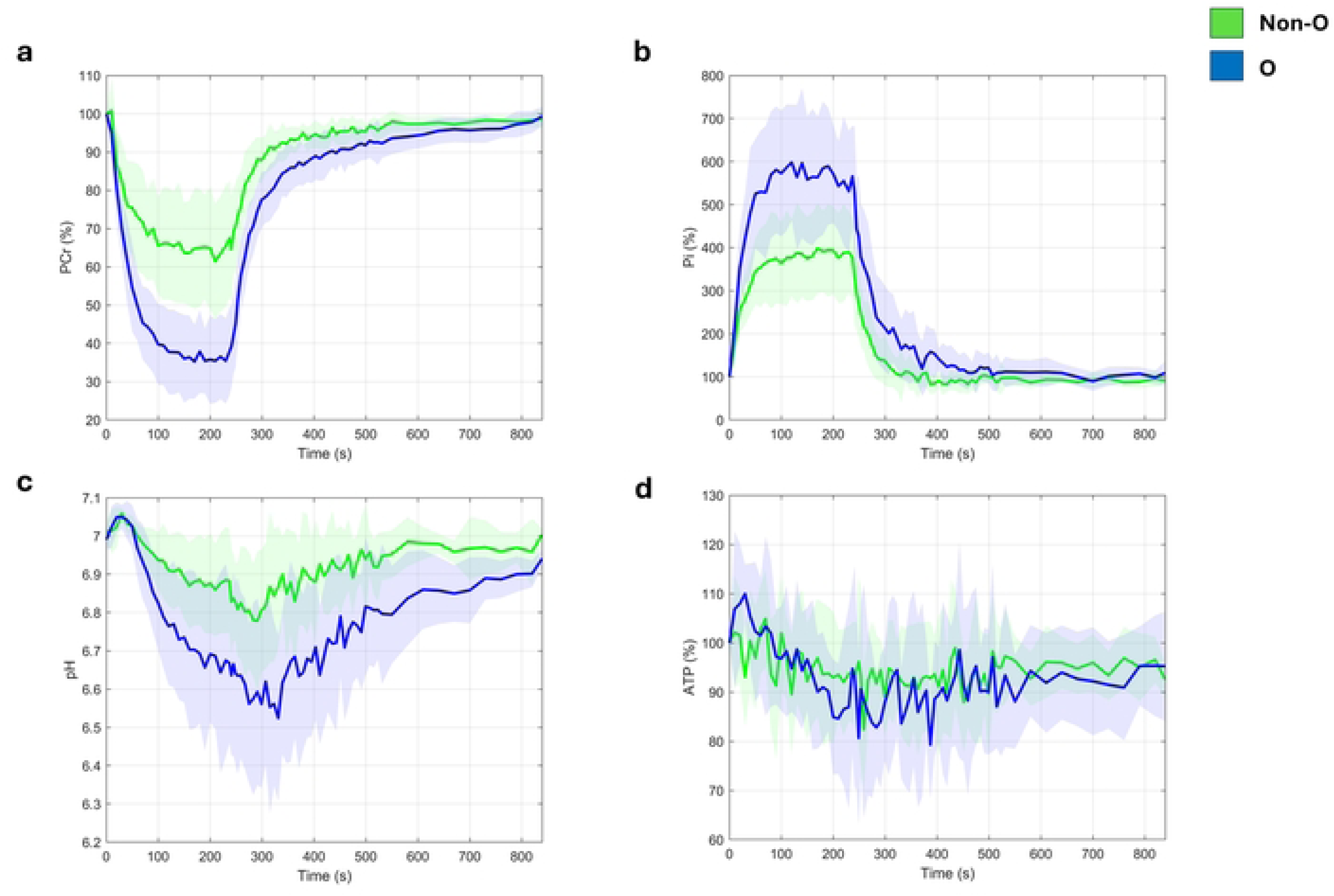
Dynamic changes in intracellular metabolites and pH during the exhaustive isotonic dorsiflexion protocol. (a) relative changes in PCr compared to rest, (b) relative changes in Pi compared to rest, (c) pH changes, and (d) relative changes in ATP compared to rest. Green lines represent non-obesity individuals (Non-O), and blue lines represent individuals with obesity (O). Data are presented as mean ± SD. PCr = phosphocreatine. Pi = inorganic phosphate (the sum of Pi1 an Pi2 is shown). ATP = adenosine triphosphate. s = seconds.

At rest, the average (±SD) SNR for PCr, Pi1, and γATP were as follows: 628 (127), 81 (13), and 94 (19) for the Non-O group, and 533 (117), 64 (13), and 66 (14) for the O group. For end-exercise values, the SNR for PCr, Pi1, Pi2, and γATP were 411 (115), 238 (35), 82 (99), and 86 (19) for Non-O, and 196 (65), 217 (88), 140 (118), and 62 (11) for O. Notably, no SNR values below 5 were observed for any individual in the given metabolites at either time point.

At rest, the percentage (±SD) of CRLB for the same peaks were 0.5 (0.1), 6.3 (1.0), and 3.0 (0.5) for Non-O, and 0.6 (0.1), 6.2 (1.1), and 3.6 (0.6) for O. At the end of exercise, the CRLB values were 2.3 (0.6), 7.4 (3.9), 32.0 (24.9), and 7.8 (1.3) for Non-O, and 4.8 (2.6), 12.4 (10.7), 17.6 (10.5), and 10.3 (3.3) for O. No CRLB values exceeded 20% for PCr or γATP in any individual, while Pi1 and Pi2 reached CRLB values above 20% at end-exercise in several subjects. Importantly, CRLB values above 20% were never observed for both Pi1 and Pi2 in the same individual at the same time point.

Outlier removal prior to spectral averaging resulted in a median (min-max) of 5 (2-17) outliers per subject for the Non-O group and 6 (1-36) outliers for the O group, with no significant difference between groups (p = 0.50, Wilcoxon rank-sum test).

The fit of PCr recovery curves, as indicated by the R² value (median [min-max]), was 0.95 (0.79-0.99) for the Non-O group and 0.98 (0.96-0.99) for the O group, with no significant difference between the groups (p = 0.18, Wilcoxon rank-sum test). All fits showed an R² value greater than 0.70.

### Correlation of muscle fatigue and ^31^P-MRS parameters

The correlations between muscle fatigue (relative drop in peak power) and the associated changes in ^31^P-MRS parameters during exercise are presented in Figure 5. Strong correlations were observed between muscle fatigue and changes in ^31^P-MRS parameters, including relative PCr drop, pH fall, the PCr recovery time constant, and the maximal Pi/PCr ratio (r ≥0.77). The goodness-of-fit (R^2^) for these relationships ranged from 0.59 to 0.68. Notably, no interaction effect of obesity was detected on the relationship between muscle fatigue and any of the changes in ^31^P-MRS parameters. All correlations were statistically significant (p-value ≤ 0.004).

**Fig 5.**
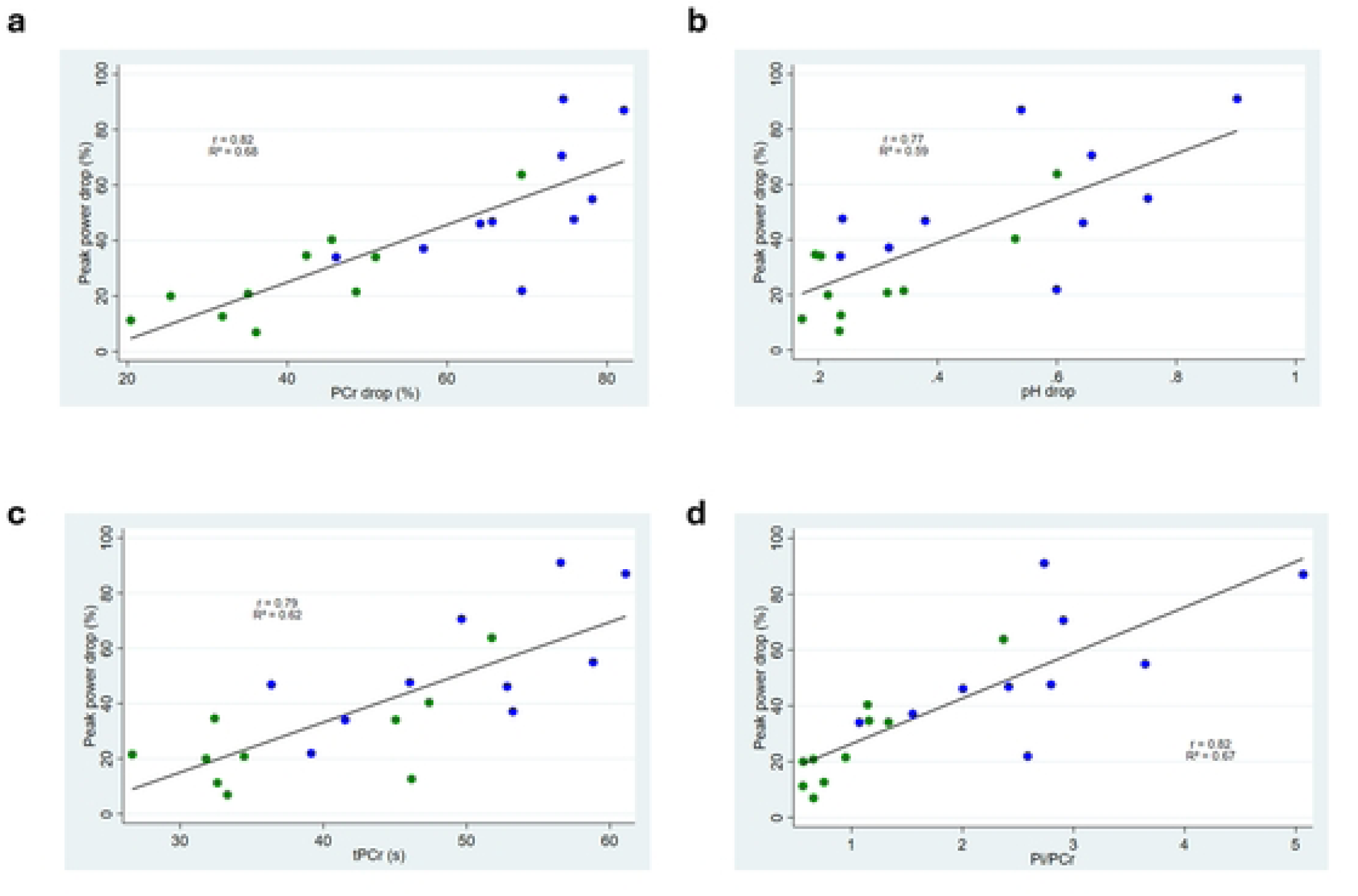
Correlations between muscle fatigue, defined as a relative drop in peak power, and ^31^P-MRS end-of-exercise parameters: (a) Relative PCr drop (P = 0.001), (b) absolute pH fall (P = 0.004), (c) PCr recovery time constant (P = 0.002), and (d) Max Pi/PCr ratio (P = 0.001). Green dots represent non-obesity individuals (Non-O), and blue dots represent individuals with obesity (O).

## Discussion

Our study aimed to validate a custom-built MR-compatible ergometer by evaluating its feasibility, reliability, and agreement with a clinically validated dynamometer for muscle contractile assessment. In-scanner dorsiflexion assessments were successfully performed with good-to-excellent reliability of muscle contractile parameters across BMI groups. Furthermore, the proof-of-concept for ^31^P-MRS in severe obesity demonstrated adequate spectral quality and consistent interrelations between muscle fatigue and ^31^P-MRS metabolic parameters. Notably, the accuracy of muscle contractile assessment on the MR ergometer and the quality of ^31^P-MRS spectra remained consistent across individuals with severe obesity and those with normal-to-overweight BMI. These results suggest that the accuracy of in-scanner muscle contractile assessments and quality of metabolic assessments in individuals with severe obesity are comparable to those without obesity, supporting the MR ergometer’s potential for clinical and research applications across diverse BMI profiles.

Clinical validation of muscle contractile examination performed in an MR environment is essential if ^31^P-MRS metabolic data interpretation is to be implemented to clinical settings. Ankle dorsiflexors has frequently been utilized to explore muscle metabolism with ^31^P-MRS [27, 37, 38]. This muscle group is critical for daily living activities, such as gait and postural control, and weakness of dorsiflexion, foot drop, is a hall mark in many disorders (e.g. brain and spinal cord disorders, as well as peripheral nerve and muscle diseases), and it plays a significant role in fall risk stratification in the elderly [39]. Voluntary activation of the dorsiflexors is feasible in most cases, making examination of this muscle group robust to challenges often encountered when assessing diverse patient groups. Therefore, the choice of ankle dorsiflexion for ^31^P-MRS studies in clinical populations appears both relevant and practical, particularly when addressing the challenges of examining individuals with obesity. This is supported by our results as all participants, regardless of BMI, successfully performed and completed the in-scanner exercise protocol.

Most tested muscle contractile parameters met the pre-set reliability criteria (ICC ≥ 0.50). However, direct comparisons with existing literature are challenging, as no prior studies have conducted a method comparison between an MR ergometer and a clinical dynamometer. Furthermore, only a limited number of studies have examined the test-retest reliability of muscle fatigue parameters in general [40–42]. While the ICCs observed in our study were slightly lower than those reported for test-retest reliability on clinical dynamometers, direct comparisons remain difficult due to methodological differences. Previous studies have predominantly assessed isokinetic contractions of the knee extensors, whereas our study focused on isotonic contractions of the ankle dorsiflexors. Notably, our findings align with intermachine reliability data for clinically validated dynamometers assessing isokinetic contractions of knee extensors and flexors (ICC: 0.41 – 0.93) [43]. Yet, the considerable variability in torque measurements across clinical dynamometers raises the question of whether they truly represent a gold standard.

Muscle fatigue measures, defined as a relative drop in muscle power or work, exhibited larger random errors than initial maximal contractile values and MVIC. This is not unexpected, given the greater complexity of quantifying muscle fatigue compared to maximal torque [44]. As a result, detecting meaningful changes in fatigue-related parameters requires larger effect sizes than those needed for assessing muscle contractile properties, consistent with prior test-retest studies [40, 45]. Interestingly, our findings suggest that muscle fatigue, assessed as a relative power drop, demonstrates higher reliability than when assessed as a decline in muscle work in individuals with severe obesity. This discrepancy may be explained by the observation that fatigue seems to disproportionately affect muscle contractile elements, with force production experiencing a smaller decline relative to muscle shortening velocity [46], an effect that appears to be more pronounced in individuals with obesity and type 2 diabetes [47, 48]. Consequently, power output may serve as a more sensitive and robust index of muscle fatigue than work in individuals with obesity.

A systematic bias was observed in measured muscle contractile parameters, with the MR ergometer underestimating initial maximum torque, maximum power, and work compared to the clinical dynamometer. This discrepancy likely arises from slight variations in subject positioning, as the practical challenges of conducting muscle contractile assessments in the MR scanner could not be entirely eliminated. However, the systematic bias remained consistent across BMI groups, and recognizing these errors allows for adjustments in future clinical interpretations.

These findings underscore the challenges of standardizing muscle contractile evaluation and emphasize the need for caution when interpreting patient progress across different devices. Nonetheless, despite differences in setup, testing environments, and a more heterogenous study population, the MR-compatible ergometer demonstrated acceptable reliability and accuracy.

Several custom-built pneumatic MR ergometers, similar to the setup proposed in our study, have been described [49–52]. Notably, pneumatic workload mechanisms have proven comparable to other workload mechanisms, such as mechanical ergometers [51]. Therefore, it seems reasonable to conclude that other existing dorsiflexion ergometers would exhibit similar agreement with clinical setups.

The quality of ^31^P-MRS spectral fitting remained robust across BMI groups, with all data points meeting SNR and CRLB criteria at rest and end-exercise. Previous studies reporting CRLB and SNR values remain limited. Sedivy et al. [51] reported a CRLB of 1.2% and an SNR of 270 for PCr at rest, while end-exercise SNR ranged from 175 to 245 depending on exercise load, with a PCr recovery goodness-of-fit (R²) of 0.83 in plantar flexors. Similarly, Xu et al. [30] presented a CRLB of 1.1% for PCr in the in vivo brain, though not directly comparable, and an R² of 0.93 for PCr recovery in dorsiflexors. Although these studies differ in the regions assessed, it is reassuring that our group with severe obesity demonstrated comparable data quality and goodness-of-fit, as no previous studies, to our knowledge, have provided detailed data on the quality of ^31^P-MRS measurements in skeletal muscles of individuals with severe obesity. Recent studies have outlined methods for outlier identification in quality control [29], but handling of outliers in ^31^P-MRS datasets remains inconsistently reported. Despite limited comparability of quality control markers across studies, the applied quality control pipeline aligns with recently published methodologies and supports the validity and consistency of ^31^P-MRS metabolite fitting in dorsiflexors across BMI groups.

This study reports significant linear relationships between muscle fatigue (relative drop in power) and muscle metabolic changes at the end of the fatiguing exercise. Specifically, our data suggest that 59% to 68% of the variance in the observed decrease in power can be explained by changes in pH, intramuscular metabolite turnover (PCr and Pi), and inherent differences in mitochondrial function. The strong associations between muscle metabolic changes and fatigue align with previous studies on ageing and muscle fatigue [22, 24, 53, 54]. Moreover, the consistency of these interrelations across groups of BMI, supports the validity of ^31^P-MRS in severe obesity. While exploring differences in fatigue and dynamic changes in intracellular metabolites between individuals with and without obesity was not the focus of our study, we observed a trend towards greater fatigue, lower oxidative capacity, lower pH, and increased PCr breakdown during exhaustive exercise in people with obesity. These findings somewhat align with recent studies suggesting lower endurance capacity in individuals with prediabetes [48], as well as greater PCr breakdown and metabolite accumulation associated with higher proportions of adipose tissue [13]. Still, the role of obesity in muscle fatigue warrants further investigation. In this context, combining metabolic measurements with accurate muscle power quantification could provide valuable insights for future exploration of this high-risk population [55].

A limitation of our study is the relatively small sample size. Despite the small sample size, the external validity of the findings is supported by the inclusion of a middle-aged, mixed-sex cohort, comprising individuals with and without obesity. Moreover, since we did not perform repeated tests with both devices, we cannot determine how much of the variation between the two methods was due to inherent variation within each device. Another limitation is the potential for familiarization effects, as even a small number of repetitive sessions of exercise may increase strength and endurance [42]. However, no order effect was apparent in this study, despite the non-randomized order of sessions.

## Conclusion

The MR-compatible ergometer demonstrated feasibility and accuracy comparable to a clinical dynamometer for assessing ankle dorsiflexor contractile function and fatigue. The reliability of muscle contractile parameters across devices was largely unaffected by BMI, and the quality of ^31^P-MRS data remained consistent across BMI groups. While a systematic bias was observed in maximum muscle torque, work, and power parameters, these differences can be accounted for in future clinical studies. The MR ergometer for dorsiflexion may therefore serve as a valuable and robust tool for evaluating muscle contractile function across individuals with varying BMI profiles, including those with severe obesity. Its ability to generate valid metabolic and reliable muscle contractile and fatigue-related data in an MR setting underscores its potential for both research and clinical applications in understanding muscle function across diverse BMI populations.

## Data Availability

A link to the 3D-printing files for essential MR ergometer components will be provided in the manuscript. The script used for analyzing muscle work and 31P-MRS data will be made publicly available. We intend to share de-identified data from human participants following acceptance.

## Acknowledgements

We sincerely thank the Technical Department at Aarhus University Hospital for their assistance in providing key components for the MR ergometer’s construction.

## Supporting information

**S1 File**. Detailed description of the MR ergometer construction

**S1 Fig 1.** Figure showing the data acquisition system for the MR ergometer

**‘S1 Fig 1. Data acquisition system for the MR ergometer.** (a) In-house analysis program (Footlogger1.1., Aarhus, Denmark), (b) Shielded cables from the strain-gauge and potentiometer, (c) Power conditioner for the strain-gauge. Filters the analog signal prior to digitalization, (d) Resistance box for the potentiometer. Receives a constant power (9v) from the power supply, (e) Data acquisition device (DAQ). The DAQ digitalizes the analog signal prior to analysis.’

**S1 Fig 2.** Figure showing the pneumatic resistance system for the MR ergometer

**‘S1 Fig 2. Pneumatic resistance system for the MR ergometer.** (a) Pneumatic tube (grey) fixed to the medical air-supply in the MR control room, (b) the pneumatic tube is fixed to the pneumatic regulator system, (c) Primary pneumatic regulator controlling the inlet-pressure, (d) Precision pressure regulator enabling fine-tuning of resistance, (e) High-precision sensor for measuring the pneumatic pressure fluctuations in the system, (f) Fixation of the pneumatic tube to the pneumatic cylinder on the MR ergometer.’

**S2 File.** Bland-Altman plots for muscle contractile outputs

**‘S2 Fig 1. Bland-Altman plots comparing muscle contractile outputs between the clinical dynamometer and MR ergometer.** Obesity corresponds to the O Group, and HC to the Non-O group. (a) MVIC (Nm). (b) Initial peak torque (Nm). (c) Initial peak power (W). (d) Initial muscle work (J). (e) Initial peak velocity (d/s). (f) Initial range of motion (ROM) (degrees). (g) Muscle fatigue (relative drop in power, %). (h) Muscle fatigue (relative drop in work, %). (i) Log-transformed MVIC. (j) Log-transformed peak velocity. In all cases, values from the clinical dynamometer were subtracted from those of the MR ergometer. The red lines represent the mean difference between modalities, while the blue lines indicate the line of equality (difference = 0). * Indicates data was considered heteroscedastic.

